# Ostial side branch stenosis in non-stenotic coronary segments: a characteristic finding in diabetes mellitus

**DOI:** 10.1101/2024.10.18.24315629

**Authors:** Yasushi Yamanaka, Yoshiharu Fujimori, Satoshi Hashimoto, Kazuto Kurihara, Tomoko Sasaki, Masayuki Yoshimura, Miki Takahashi, Tadamasa Wakabayashi, Taku Imai

**Affiliations:** The Department of Cardiology, Suwa Central Hospital, Chino, Nagano, Japan

## Abstract

2.

**Background:** Ostial stenosis, a narrowing at the origin of coronary side branches, is commonly observed in branches diverging from stenotic coronary segments. However, it may also occur in branches diverging from non-stenotic segments, and the factors contributing to this phenomenon remain unclear.

**Objectives:** This study aimed to assess the incidence of ostial stenosis in coronary side branches diverging from non-stenotic segments and to identify associated risk factors.

**Methods:** We conducted a retrospective analysis of first-time, elective coronary angiograms (CAGs) from 884 patients. Side branches with a diameter larger than 2mm were included. Ostial stenosis was defined by two criteria: 1) the side branch diverged from a coronary segment with less than 25% diameter stenosis, and 2) the side branch exhibited ostial narrowing with more than 50% diameter stenosis. Clinical factors, including diabetes mellitus and lipid profiles, were assessed. Multivariate logistic regression was used to identify independent predictors of ostial stenosis.

**Results:** Of the 4,739 side branches analyzed, 508 (10.7%) exhibited ostial stenosis. Ostial stenosis was present in 285 (32.2%) patients. Patients with ostial stenosis had a significantly higher prevalence of diabetes mellitus (60.0% vs. 33.0%, *p*<0.00001) and lower high-density lipoprotein (HDL) cholesterol levels (48.5 ± 14.6 mg/dL vs. 52.0 ± 14.9 mg/dL, *p*<0.01) compared to those without stenosis. No significant differences were found in other clinical factors. Multivariate analysis identified diabetes mellitus as an independent predictor of ostial stenosis (odds ratio: 0.29, 95% CI: 0.20-0.43, *p*<0.0001).

**Conclusions:** Ostial stenosis in side branches diverging from non-stenotic coronary segments is significantly associated with diabetes mellitus. In diabetic patients, factors such as negative arterial remodeling, insulin resistance, and endothelial dysfunction likely play key roles in the development of this condition. These findings suggest that ostial stenosis could be an additional marker of advanced coronary artery disease in diabetes.

## 3. Introduction

Percutaneous coronary intervention (PCI) is widely recognized as an effective treatment for coronary artery disease (CAD).^1,2^ While standard stent implantation effectively addresses typical stenotic lesions in the main coronary arteries, bifurcation lesions, particularly ostial narrowing of a side branch, present unique challenges.^3-5^ Ostial stenosis is commonly observed in side branches diverging from stenotic coronary segments, where plaque at the side branch ostium is often considered part of the plaque burden from the main coronary artery. However, ostial stenosis can also occur in side branches diverging from non-stenotic coronary segments, a phenomenon that is less well-understood and less commonly studied.

Coronary arteriosclerosis progresses rapidly in patients with CAD risk factors, with diabetes mellitus being one of the most significant contributors.^6^ In patients with diabetes mellitus, arteriosclerosis is accelerated by mechanisms such as insulin resistance and endothelial dysfunction.^7-11^ On coronary angiography (CAG), diabetic patients often exhibit diffuse stenotic lesions, calcification, and multivessel disease.^12,13^ While diabetes mellitus is known to promote the development of stenosis in the main coronary arteries^14^, its effects on side branches, particularly in non-stenotic coronary segments, have not been as thoroughly examined.

### Study objective

The objective of this study was to determine the incidence of ostial stenosis in side branches diverging from non-stenotic coronary segments, and to investigate the underlying factors, with a particular focus on the role of diabetes mellitus.

## 4. Methods

### Study design and participants

This retrospective study analyzed first-time, elective diagnostic CAGs performed at Suwa Central Hospital between 2009 and 2019. The study aimed to investigate the relationship between the presence of ostial stenosis and background factors such as diabetes mellitus, age, and gender. Patients included in the study had no prior coronary interventions or surgical coronary therapies, and were excluded those with acute coronary syndromes, including unstable angina or acute myocardial infarction. Additionally, CAGs were excluded if a guidewire was used for distal coronary pressure assessment, regardless of whether PCI was performed.

### Primary outcome and inclusion criteria

The primary outcome of this study was defined as ostial stenosis, characterized by narrowing at the origin of coronary side branches. Ostial stenosis was defined by two criteria: 1) the side branch diverging from a coronary segment with less than 25% diameter stenosis, and 2) the side branch exhibiting ostial narrowing with more than 50% diameter stenosis. Additionally, side branches included in the analysis were those with a diameter greater than 2 mm, while significant stenosis in the main coronary artery was defined as greater than 75% diameter stenosis.

### Procedure

CAG was performed via radial or femoral artery access using 4- or 5-Fr diagnostic catheters with non-ionic contrast media (Iopamiron®; Bracco, Milan, Italy). An intravenous bolus of heparin (100 IU/kg) was administered prior to the procedure, and intracoronary nitrates were used to prevent vasospasm. Five experienced cardiologists independently assessed the severity of coronary stenosis via visual inspection.

### Laboratory Data

Data were obtained from medical records, providing comprehensive clinical information relevant to the study measured using standard clinical methods. The analysis included age, sex, BMI, and risk factors such as hypertension, diabetes, dyslipidemia, and smoking history as variables of interest. Biomarkers such as HDL cholesterol level, triglyceride (TG) level, LDL cholesterol level, and HbA1c were also adopted.

### Statistical analysis

Continuous variables were summarized as means and standard deviations, and categorical variables as percentages. Comparisons between groups were performed using the unpaired t-test for continuous variables and the chi-square test for categorical variables. Multivariate logistic regression analysis was employed to identify independent predictors of ostial stenosis, adjusting for potential confounders. Variables with a *p*-value <0.05 in univariate analysis were included in the multivariate model. Subgroup analyses were conducted to explore differences among specific populations, such as age groups or sex, to identify potential variations in the effects of predictors. Sensitivity analyses were conducted to assess the robustness of the findings, including the exclusion of patients with extreme HDL cholesterol levels (<20 mg/dL or >80 mg/dL). Missing data were minimal and handled using pairwise deletion to ensure maximum use of available data for each analysis. All statistical analyses were performed using Excel (Microsoft, Redmond, WA, USA), MedCalc (MedCalc Software, Mariakerke, Belgium), and SPSS version 23.0 (IBM Japan, Ltd, Tokyo, Japan). A *p*-value <0.05 was considered statistically significant.

## 5. Results

### Participant characteristics and inclusion

A total of 884 CAGs from 884 patients were retrospectively included and analyzed in this study. Figure 1A demonstrates ostial stenosis (90% diameter stenosis) in a side branch diverging from a non-stenotic coronary segment, while Figure 1B shows ostial stenosis (90% diameter stenosis) from a stenotic coronary segment. The former type was included in the definition of ostial stenosis.

**Figure 1.**
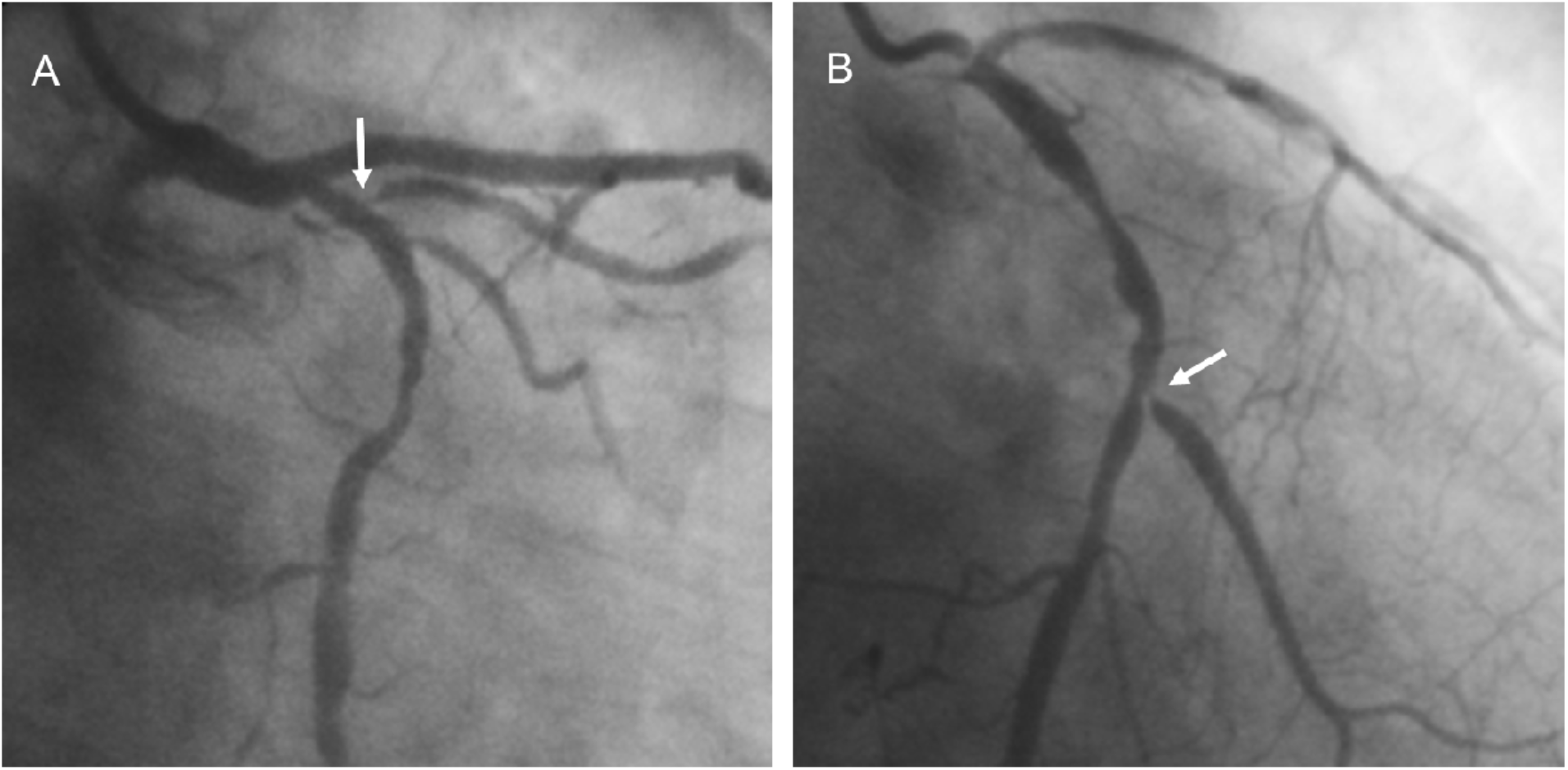
Representative images of ostial stenosis in coronary side branches. **A:** Ostial stenosis in a side branch diverging from a non-stenotic coronary segment **(arrow)**. **B:** Ostial stenosis in a side branch diverging from a stenotic coronary segment **(arrow)**.

There were no significant differences in baseline characteristics between patients with and without ostial stenosis, such as age, gender, or body mass index (BMI)(Table1). However, diabetes mellitus was significantly more prevalent in the ostial stenosis group (59.6%) compared to the non-stenosis group (33.0%, *p*<0.00001). Additionally, HDL cholesterol levels were significantly lower in the ostial stenosis group (48.5±14.6 mg/dL) compared to the non-stenosis group (52.0±14.9 mg/dL, *p*<0.01), and hemoglobin A1c (HbA1c) was significantly higher in the stenosis group (6.7±1.6% vs.6.2±1.2%, *p*<0.00001). Missing data for key variables (e.g., diabetes status and HDL cholesterol levels) were minimal, affecting less than 5% of participants.

**Table 1.** Baseline characteristics of patients with and without ostial stenosis.

| Variable | Ostial stenosis +<br>(n=285) | Ostial stenosis –<br>(n=599) | *p*-value |
| --- | --- | --- | --- |
| **Patients (n=884), n (%)** | 285 (32.2) | 599 (67.8) | - |
| **Age, (mean ± SD)** | 67.6 ± 10.2 | 67.2 ± 11.5 | >0.05 |
| **Male, n (%)** | 221 (77.5) | 480 (80.1) | >0.05 |
| **BMI, (mean ± SD)** | 23.5 ± 3.6 | 23.2 ± 3.8 | >0.05 |
| **Risk factors** |  |  |  |
| Hypertension, n (%) | 159 (55.7) | 328 (54.7) | >0.05 |
| Diabetes mellitus, n (%) | 170 (59.6) | 201 (33.0) | <0.00001 |
| Dyslipidemia, n (%) | 135 (47.4) | 275 (45.9) | >0.05 |
| History of smoking, n (%) | 106 (37.2) | 195 (32.6) | >0.05 |
| **Blood chemistry** |  |  |  |
| HDL cholesterol (mg/dL, mean ± SD) | 48.5 ± 14.6 | 52.0 ± 14.9 | <0.01 |
| LDL cholesterol (mg/dL, mean ± SD) | 122.7 ± 80.2 | 115.8 ± 38.0 | >0.05 |
| Triglyceride (mg/dL, mean ± SD) | 139.3 ± 88.1 | 142.3 ± 97.9 | >0.05 |
| HbA1c (% , mean ± SD) | 6.7 ± 1.6 | 6.2 ± 1.2 | <0.00001 |

BMI, body mass index; SD, standard deviation; HDL, high-density lipoprotein; LDL, low-density lipoprotein; HbA1c, hemoglobin A1c; ns, not significant.

### Coronary angiograms and coronary arteriosclerosis findings

Table 2 summarizes the CAG results. Ostial stenosis was observed in 285 cases (32.2%) of the 884 CAGs. Multivessel disease was significantly more frequent in the ostial stenosis group compared to the non-stenosis group (30.2% vs. 14.4%, *p*<0.00001).

**Table 2.**
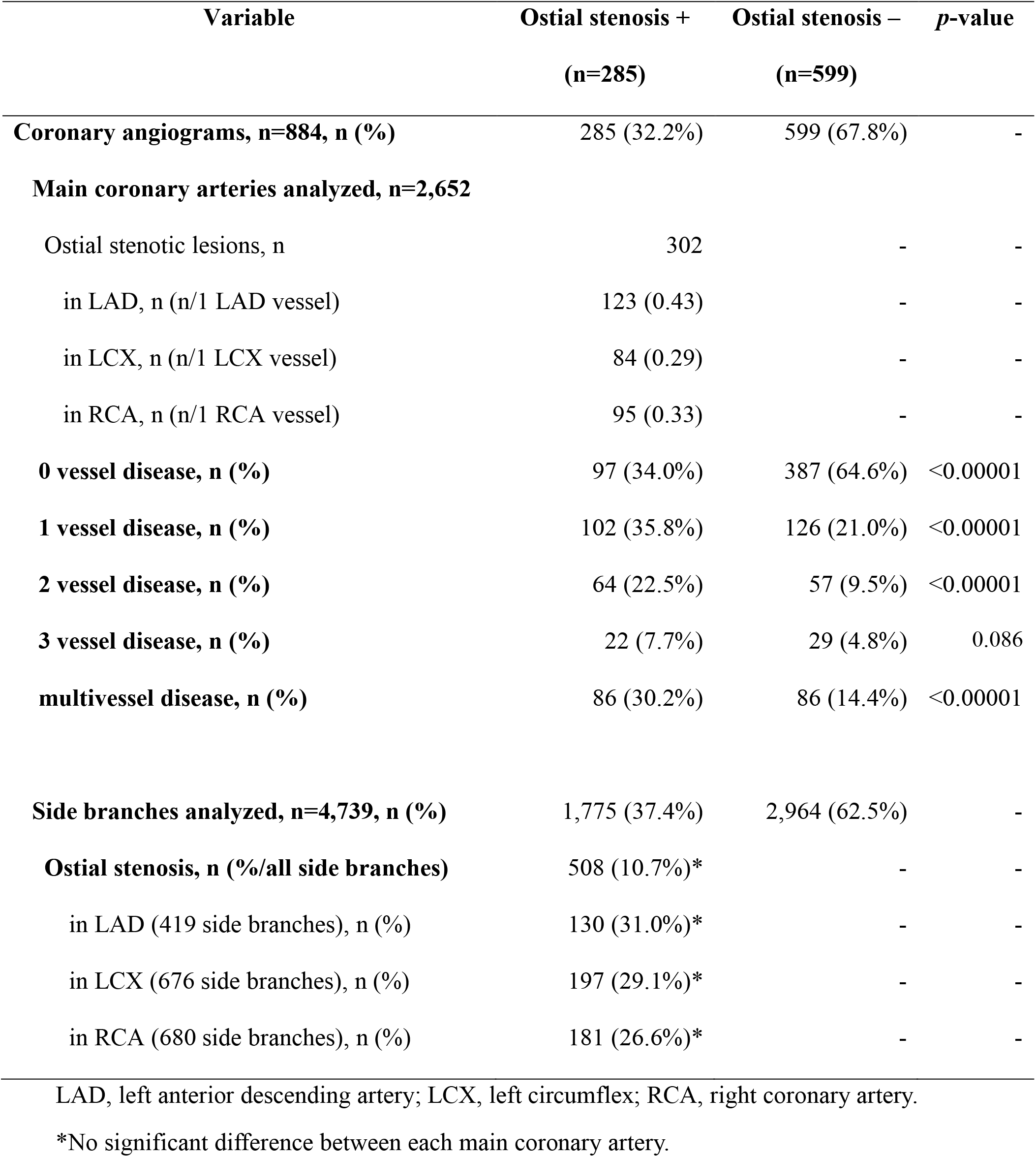
CAG data of patients with and without ostial stenosis.

### Main findings and specific analysis of side branches

Of the 4,739 side branches analyzed, 508 (10.7%) exhibited ostial stenosis. Ostial stenosis occurred in 130 (31.0%) of 419 side branches in the left anterior descending (LAD) artery, 197 (29.1%) of 676 side branches of the left circumflex (LCX) artery, and 181 (26.6%) of 695 side branches of the right coronary artery (RCA), with no significant differences in prevalence across the arteries.

### Multivariate logistic regression analysis

Multivariate logistic regression analysis was conducted to identify independent predictors of ostial stenosis. Only age, sex, and the four risk factors (hypertension, diabetes, dyslipidemia, and smoking history) were included, totaling six variables in the model. The analysis identified diabetes mellitus as the sole independent predictor of ostial stenosis, with an odds ratio (OR) of 0.29 (95% CI: 0.20-0.43, *p*<0.0001) after adjusting for other variables (Table3).

**Table 3.** Multivariate logistic regression analysis for ostial stenosis.

| **Variable** | **Odds ratio** | **95%CI** | ***p*-value** |
| --- | --- | --- | --- |
| Age | 0.98 | 0.96-1.00 | 0.037 |
| Sex (male) | 0.91 | 0.59-1.38 | 0.65 |
| Hypertension | 1.03 | 0.69-1.53 | 0.88 |
| Diabetes mellitus | 0.29 | 0.20-0.43 | <0.0001 |
| Dyslipidemia | 1.03 | 0.70-1.52 | 0.87 |
| History of smoking | 1.25 | 0.79-2.00 | 0.35 |

## 6. Discussion

This study demonstrated that ostial stenosis in side branches diverging from non-stenotic coronary segments is associated with more advanced arteriosclerosis, a higher incidence of stenotic lesions, and a greater prevalence of multivessel disease. Diabetes mellitus was identified as an independent risk factor for ostial stenosis in these branches.

While previous studies have shown that bifurcation lesions, including ostial stenosis, are more common in diabetic patients^15^, the condition of the main coronary segment from which the side branch diverges has been largely overlooked. Our findings highlight that ostial stenosis in side branches from non-stenotic segments is a notable feature of diabetic patients.

In diabetic coronary arteries, diffuse stenotic lesions, calcification, and multivessel disease are well-established characteristics.^12,13^ This study adds ostial stenosis of side branches diverging from non-stenotic segments as another important feature. The incidence of ostial stenosis was around 10% in the side branches analyzed, with a slight prevalence in the LAD, although this was not statistically significant.

The mechanism underlying ostial stenosis in side branches diverging from non-stenotic coronary segments is not fully understood, but reduced shear stress and endothelial dysfunction likely play a role. Sheer stress is typically lower in the upstream ostial area, contributing to impaired endothelial function and accelerated plaque formation.^16-19^ Our findings suggest that diabetes mellitus, through mechanisms such as insulin resistance and endothelial dysfunction, contributes significantly to the development of ostial stenosis.

We propose that negative arterial remodelling in diabetes mellitus^20-22^ plays a key role in the development of ostial stenosis. In non-diabetic coronary arteries, positive remodelling allows for arterial expansion, preventing ostial narrowing. However, in diabetic arteries, negative remodelling leads to centripetal growth of the intimal tissue, resulting in stenosis (Figure 2).

**Figure 2.**
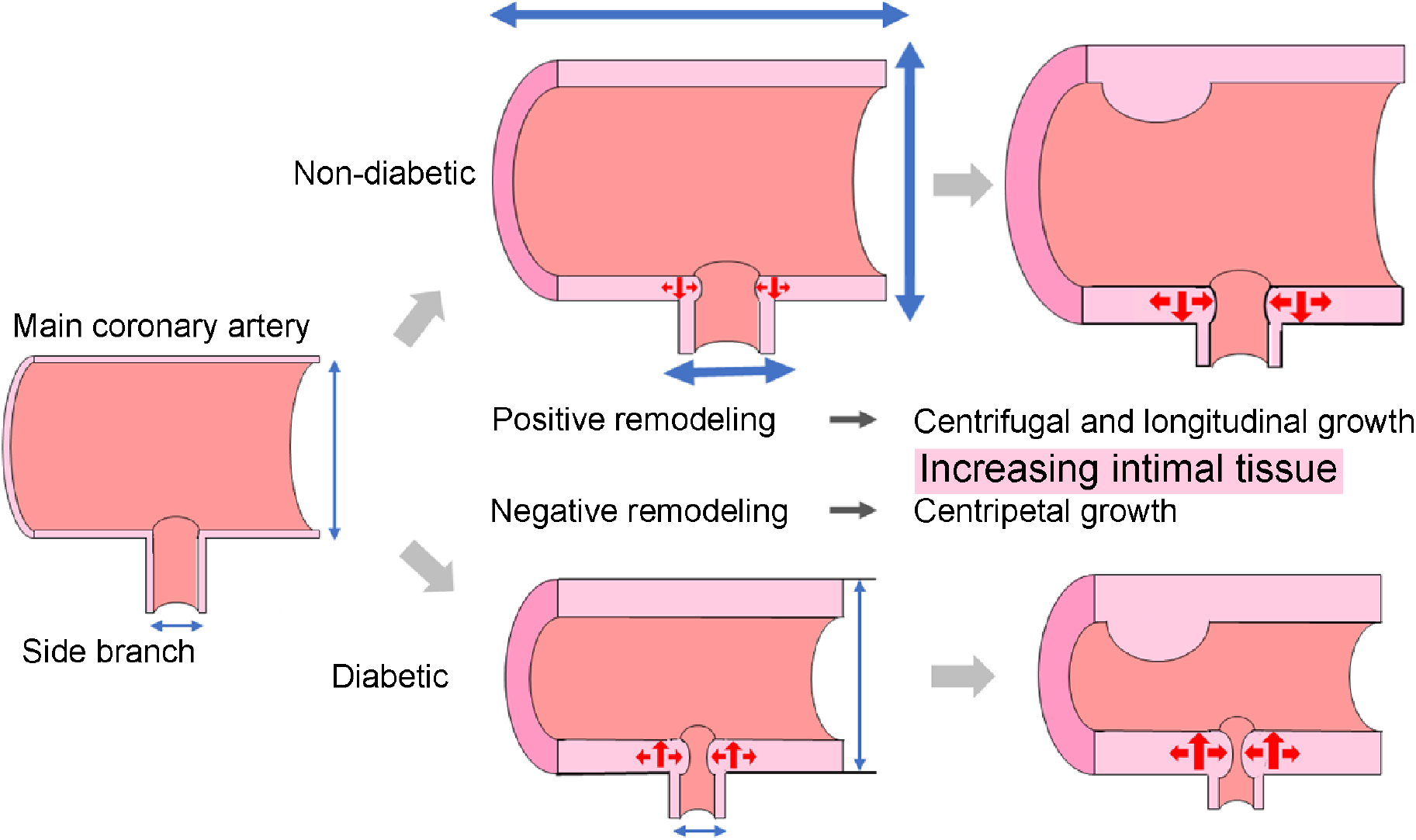
Possible mechanisms of the progression of ostial stenosis in diabetes mellitus. In positive remodeling in non-diabetic individuals, intimal tissue grows centrifugally and longitudinally**(upper panel)**. Incontrast, under negative remodeling in diabetes mellitus, intimal tissue grows centripetally**(lower panel)**.

Ostial stenosis should be considered an additional characteristic of diabetic coronary artery disease, alongside diffuse lesions, calcification, and multivessel disease.

### Study limitation

While the number of enrolled CAGs was sufficient for robust analysis, there are several limitations. First, stenosis assessment was based on visual estimation by cardiologists, which may introduce variability. Although this method is widely accepted in clinical practice, automated measurement tools might offer more precision in future studies. Second, this study was limited to patients undergoing elective CAG, which may not represent the broader population with different clinical presentations. Lastly, potential confounders such as other forms of arteriosclerosis, medication, and lifestyle factors were not fully captured in the dataset.

## Conclusions

Ostial stenosis in side branches diverging from non-stenotic coronary segments is a significant finding in patients with coronary artery disease, especially those with diabetes mellitus. Identifying diabetes mellitus as a primary risk factor for this type of stenosis enhances our understanding of coronary artery disease’s progression in diabetic patients.

## Data Availability

All data supporting the findings of this study are included within the manuscript.

## Abbreviations

CAG: coronary angiogram
PCI: percutaneous coronary intervention
HDL: high-density lipoprotein
CAD: coronary artery disease

## 7. Acknowledgements

None.

